# COVID-19 rebound after Paxlovid treatment during Omicron BA.5 vs BA.2.12.1 subvariant predominance period

**DOI:** 10.1101/2022.08.04.22278450

**Authors:** Lindsey Wang, Nora D. Volkow, Pamela B. Davis, Nathan A. Berger, David C. Kaelber, Rong Xu

**Author notes:** Corresponding Author: Rong Xu, PhD, Center for Artificial Intelligence in Drug Discovery, Case Western Reserve University, 10900 Euclid Avenue, Cleveland, OH 44106.

## Abstract

Paxlovid was authorized by FDA to treat mild-to-moderate COVID-19. In May 2022, the Centers for Disease Control and Prevention (CDC) issued a Health Alert Network Health Advisory on potential COVID-19 rebound after Paxlovid treatment. Since June 2022, Omicron BA.5 has become the dominant subvariant in the US, which is more resistant to neutralizing antibodies than the previous subvariant BA.2.12.1. Questions remain as to how COVID-19 rebound after Paxlovid treatment differs between the BA.5 and BA.2.12.1 subvariants. This is a retrospective cohort study of 15,913 patients who contracted COVID-19 between 5/8/2022-7/18/2022 and were prescribed Paxlovid within 5 days of their COVID-19 infection. The study population was divided into 2 cohorts: (1) BA.5 cohort (n=5,161) – contracted COVID-19 during 6/19/22-7/18/22 when BA.5 was the predominant subvariant^2^. (2) BA.2.12.1 cohort (n=10,752) – contracted COVID-19 during 5/8/22-6/18/22 when the BA.2.12.1 was the predominant subvariant. The risks of both COVID-19 rebound infections and symptoms 2-8 days after Paxlovid treatment were higher in the BA.5 cohort than in the propensity-score matched BA.2.12.1 cohort: rebound infections (Hazard Ratio or HR: 1.32, 95% CI: 1.06-1.66), rebound symptoms (HR: 1.32, 95% CI: 1.04-1.68). As SARS-CoV-2 evolves with successive subvariants more evasive to antibodies, continuous vigilant monitoring is necessary for COVID-19 rebounds after Paxlovid treatment and longer time duration of Paxlovid treatment warrants evaluation.

## Background

Paxlovid was authorized by FDA to treat mild-to-moderate COVID-19. In May 2022, the Centers for Disease Control and Prevention (CDC) issued a Health Alert Network Health Advisory on potential COVID-19 rebound after Paxlovid treatment^1^. Since June 2022, Omicron BA.5 has become the dominant subvariant in the US^2^. It is more resistant to neutralizing antibodies than the previous subvariant BA.2.12.1^3^. Questions remain as to how COVID-19 rebound after Paxlovid treatment differs between the BA.5 and BA.2.12.1 subvariants.

## Objective

To compare COVID-19 rebound risks after Paxlovid treatments in patients who contracted COVID-19 during the BA.5 and BA.2.12.1 subvariant predominance period in the US.

## Methods

We used the TriNetX Analytics COVID-19 Research Network platform that contains nation-wide and real-time de-identified electronic health records (EHRs) of 98 million unique patients from 76 health care organizations with both inpatient and outpatient facilities across 50 states in the US^4^, covering diverse geographic locations, age groups, racial and ethnic groups, income levels and insurance types. TriNetX Analytics Platform performs statistical analyses on patient-level data and only reports on population level data and results without including protected health information (PHI) identifiers. MetroHealth System, Cleveland OH, Institutional Review Board determined that research using TriNetX is not Human Subject Research and therefore exempt from review. The study population comprised 15,913 patients age ≥ 12 years who contracted COVID-19 between 5/8/2022-7/18/2022 and were prescribed Paxlovid within 5 days of their COVID-19 infection. The study population was divided into 2 cohorts: (1) BA.5 cohort (n=5,161) – contracted COVID-19 during 6/19/22-7/18/22 when BA.5 was the predominant subvariant^2^. (2) BA.2.12.1 cohort (n=10,752) – contracted COVID-19 during 5/8/22-6/18/22 when the BA.2.12.1 was the predominant subvariant^2^.

We compared risks for COVID-19 rebound outcomes in BA.5 and BA.2.12.1 cohorts before and after propensity-score matching (1:1 matched using a nearest neighbor greedy algorithm with a caliper of 0.25 times the standard deviation) for covariates that are related to COVID-19 infections and outcomes^5^ (**Table 1** and **eMethod)**. Rebound COVID-19 infections and symptoms (details in **eMethod**) during 2 to 8 days after Paxlovid treatment were followed and Kaplan-Meier survival analysis performed to estimate probability of outcomes. Cox’s proportional hazards model was used to compare the two cohorts. Hazard ratio (HR) and 95% confidence intervals were used to describe the relative hazard of rebound outcomes based on comparison of time to event rates. All statistical tests were conducted within the TriNetX Advanced Analytics Platform on 8/4/2022 using R’s Survival package, version 3.2-3. Details of the TriNetX database, covariates, propensity-score matching are in **eMethods**.

**Table 1.**
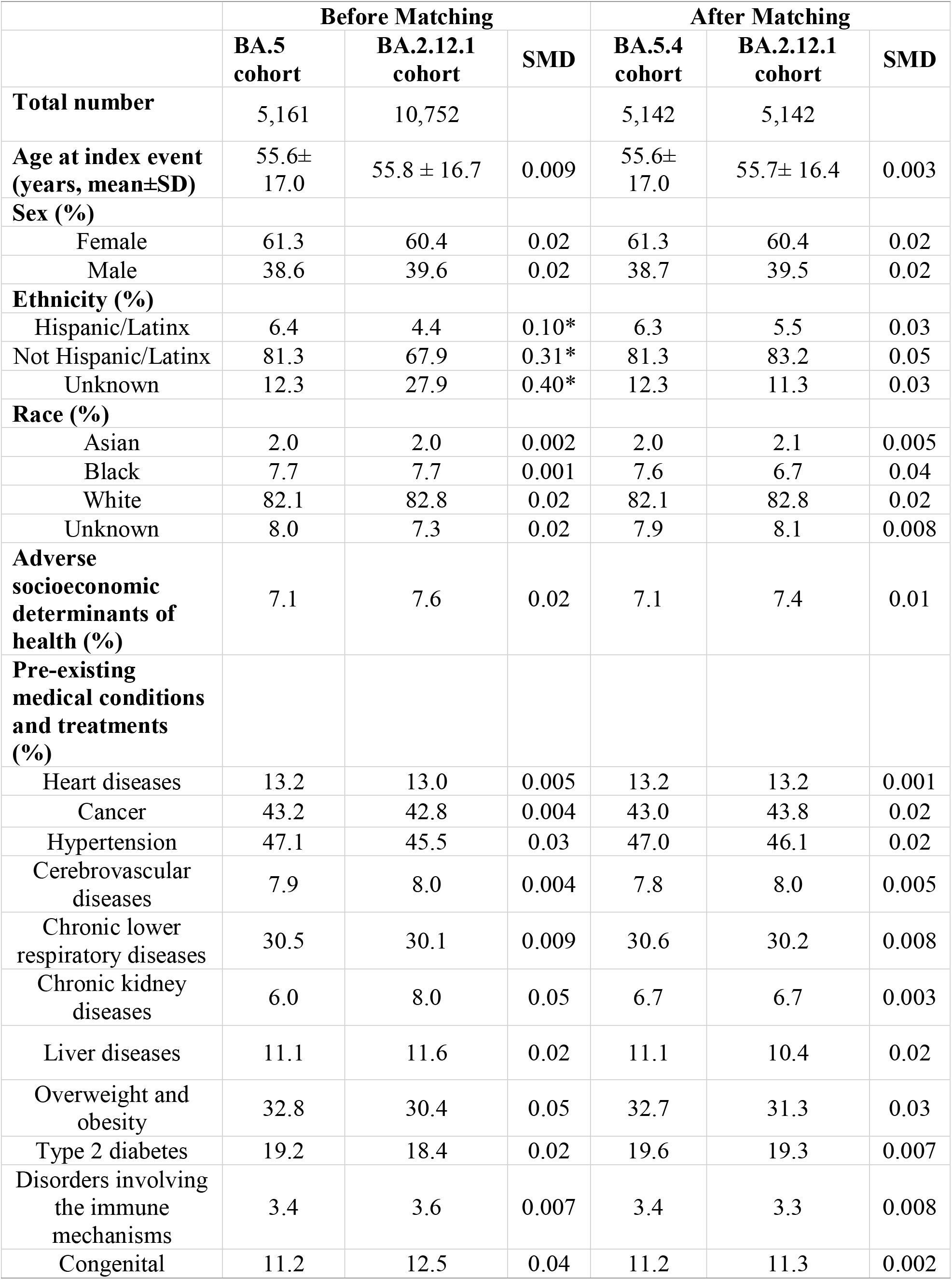
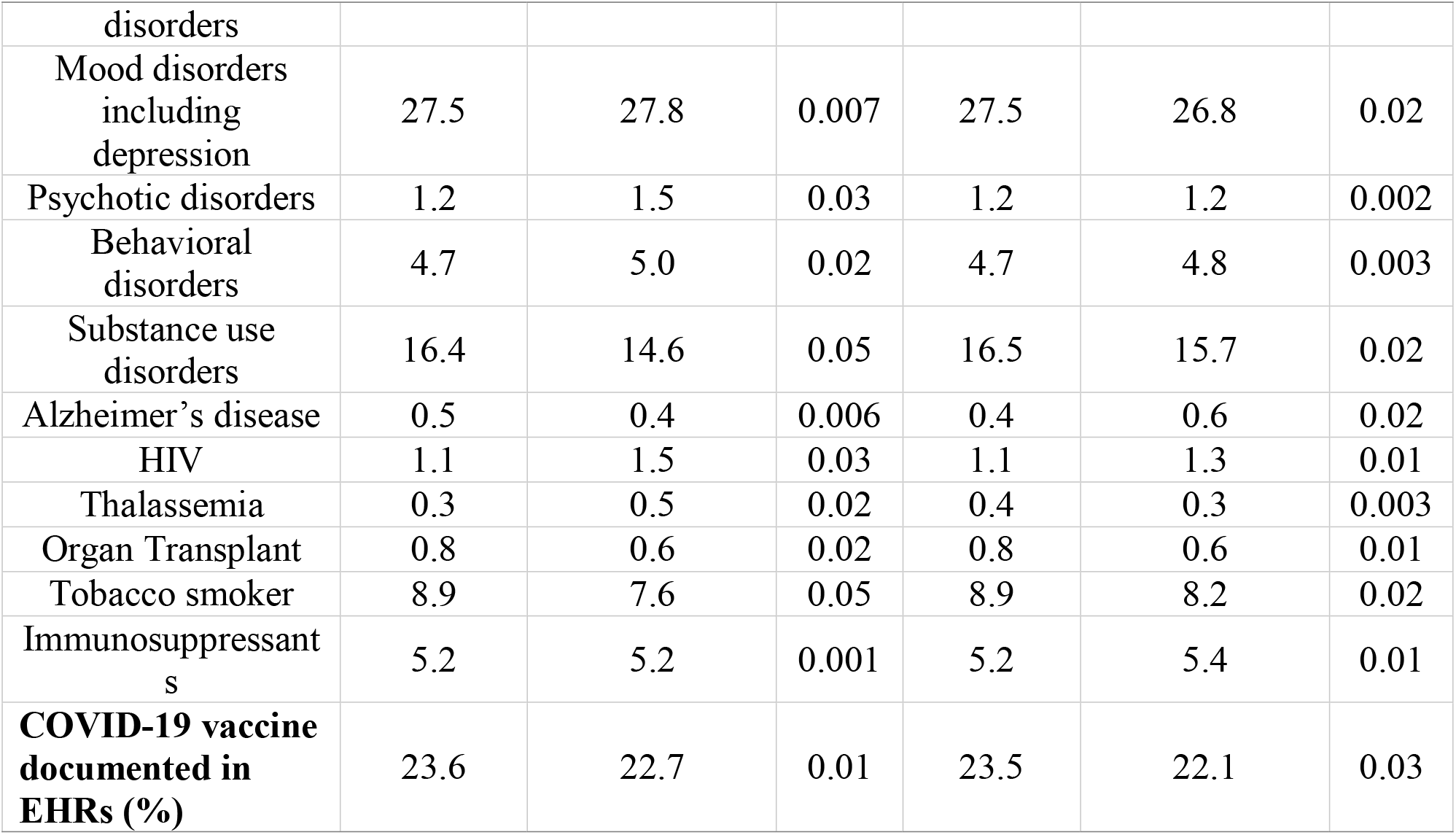
Characteristics of the study population before and after propensity-score matching (1:1 matching based on greedy nearest-neighbour matching with a caliper of 0.25 x standard deviation). BA.5 cohort – who contracted COVID-19 during 6/19/22-7/15/22 when the BA.5 was the predominant subvariant in the US and were prescribed Paxlovid within 5 days of their COVID-19 infection. BA.2.12.1 cohort – who contracted COVID-19 during 5/8/22-6/18/22 when the BA.2.12.1 was the predominant subvariant and were prescribed Paxlovid within 5 days of their COVID-19 infection. The status for adverse socioeconomic determinants of health, medical conditions, immunosuppressant usage, organ transplants, and EHR-based COVID-19 vaccination status were based on presences of related codes in patients’ EHRs anytime up to 1 day before Paxlovid treatment. SMD – standardized mean differences. *SMD greater than 0.1, a threshold being recommended for declaring imbalance.

## Findings

The BA.5 and BA.2.12.1 cohorts did not differ except that the BA.5 cohort comprised more Hispanics. After propensity-score matching, the two cohorts were balanced (**Table 1**). The cumulative risk of COVID-19 rebound infection 2-8 days after Paxlovid treatment was 2.81% and 3.42% in the BA.5 cohort and the BA.2.12.1 cohort respectively. The instantaneous risk of rebound symptoms was higher in the BA.5 cohort than in the BA.2.12.1cohort (HR: 1.24, 95% CI: 1.01-1.53) but did not differ for rebound infections. After propensity-score matching, instantaneous risks of both rebound infections and symptoms were higher in the BA.5 cohort than in the matched BA.2.12.1 cohort: rebound infections (HR: 1.32, 95% CI: 1.06-1.66), rebound symptoms (HR: 1.32, 95% CI: 1.04-1.68) (**Figure 1**).

**Figure 1.**
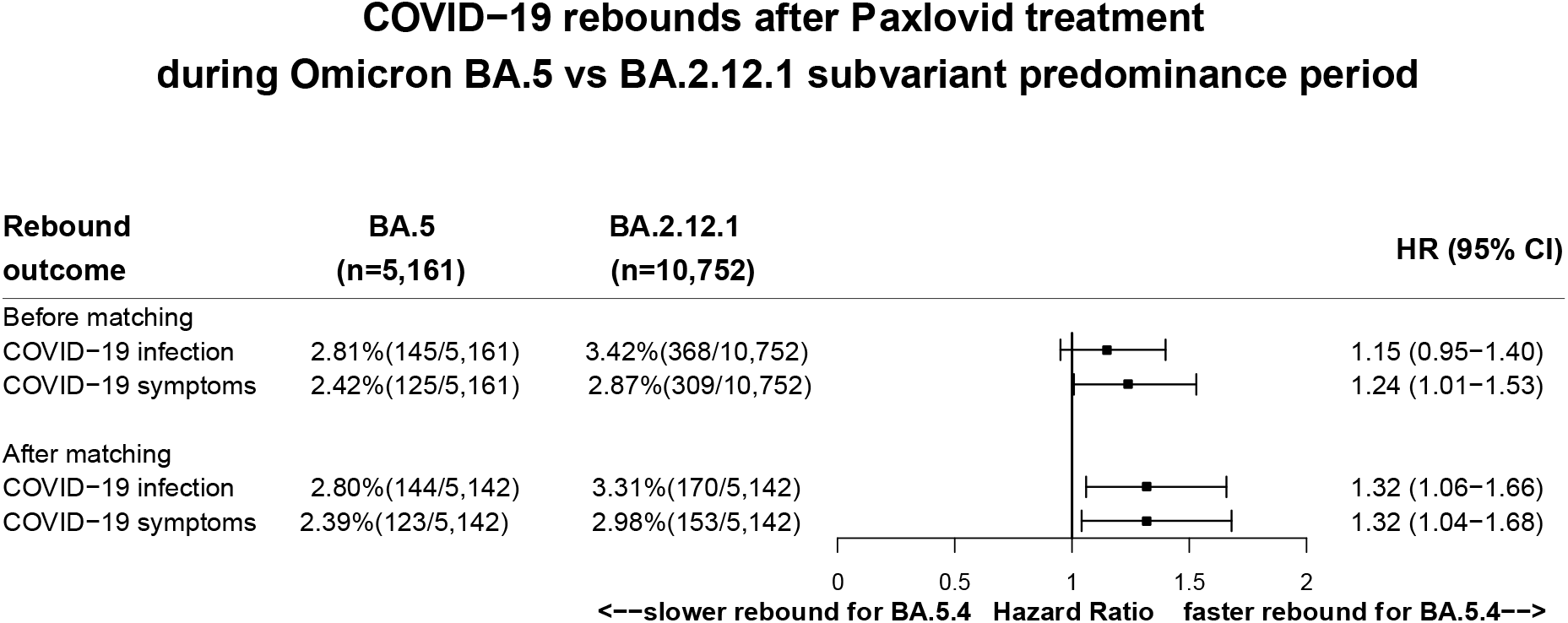
COVID-19 rebounds 2-8 days after Paxlovid treatment between the BA.5 and BA.2.12.1 cohorts before and after propensity-score matching.

## Discussion

The probability of COVID-19 rebound 2-8 days after Paxlovid treatment was higher in patients who contracted COVID-19 during the BA.5 than the BA.2.12.1 predominance period. As SARS-CoV-2 evolves with successive subvariants more evasive to antibodies, continuous vigilant monitoring is necessary for COVID-19 rebounds after Paxlovid treatment and longer time duration of Paxlovid treatment may be evaluated. Our study has several limitations: First, patient EHRs may be subject to over/mis/under-diagnosis and unmeasured confounders such as compliance with medication adherence and completion of treatment regimes. However, our comparison populations were both drawn from the TriNetX dataset, therefore these issues should not substantially affect the relative risk analyses. Second, patients in the TriNetX database may not necessarily represent the entire US population. Also, the accumulative risks of COVID-19 rebound documented in EHRs could be a substantial under-estimate of true prevalence, though this under-count should not substantially affect the comparison of instantaneous risks between the BA.5 and BA.2.12.1 cohorts. Nonetheless, results from our study need to be validated in other populations.

## Supporting information

eMethod

## Data Availability

All data produced in the present work are contained in the manuscript

## Data Availability

All data produced in the present work are contained in the manuscript

## Contributors

RX conceived and designed the study and drafted the manuscript. LW performed data analysis and prepared tables and figures and participated in manuscript preparation. NDV, NAB, PBD, DCK critically contributed to study design, result interpretation and manuscript preparation. We confirm the originality of content. RX had full access to all the data in the study and takes responsibility for the integrity of the data and the accuracy of the data analysis.

## Acknowledgments

We acknowledge support from National Institute on Aging (grants nos. AG057557, AG061388, AG062272, AG076649), National Institute on Alcohol Abuse and Alcoholism (grant no. AA029831), the Clinical and Translational Science Collaborative (CTSC) of Cleveland (grant no. TR002548), National Cancer Institute Case Comprehensive Cancer Center (CA221718, CA043703, CA2332216).

## Meeting Presentation

No

