## Supplementary material for "COVID-19 rebound after Paxlovid treatment during Omicron BA.5 vs BA.2.12.1 subvariant predominance period": eMethod

**Description of TriNetX database**

The data used in this study was collected on August 4, 2022 from the TriNetX COVID-19 Research Network, which provided access to electronic medical records (diagnoses, procedures, medications, laboratory values, genomic information) from over 98 million patients from 76 healthcare organizations. TriNetX, LLC is compliant with the Health Insurance Portability and Accountability Act (HIPAA). Any data displayed on the TriNetX Platform in aggregate form, or any patient level data provided in a data set generated by the TriNetX Platform, only contains de-identified data as per the de-identification standard defined in Section §164.514(a) of the HIPAA Privacy Rule. MetroHealth System, Cleveland, Ohio, IRB has determined research using TriNetX, is not Human Subject Research and therefore exempt from IRB review.

TriNetX is a platform that de-identifies and aggregates electronic health record (EHR) data from contributing healthcare systems, most of which are large academic medical institutions with both inpatient and outpatient facilities at multiple locations, across 50 states in the US. TriNetX Analytics provides web-based and secure access to patient EHR data from hospitals, primary care, and specialty treatment providers, covering diverse geographic locations, age groups, racial and ethnic groups, income levels and insurance types including various commercial insurances, governmental insurance (Medicare and Medicaid), self-pay/uninsured, worker compensation insurance, military/VA insurance among others.

Self-reported race and ethnicity data in TriNetX comes from the underlying clinical EHR systems of the contributing healthcare systems. TriNetX maps race and ethnicity data from the contributing healthcare systems to the following categories: (1) Race: Asian, American Indian or Alaskan Native, Black or African American, Native Hawaiian or Other, White, Unknown race; and (2) Ethnicity: Hispanic or Latino, Not Hispanic or Latino, Unknown Ethnicity.

**Covariates, outcome measures, propensity-score matching**

**eTable 1**: Covariates used for propensity-score matching, their standardized names, codes, data types and time window that are used in the TriNetX database.

| **Covariates** | **Name, code** | **Data type** | **Time Window** |
| --- | --- | --- | --- |
| **Demographics** |  |  |  |
| Age at Index | Age at Index | continuous | Age at Paxlovid treatment |
| Female | F | present/absent |  |
| Male | M | present/absent |  |
| Hispanic/Latino | Hispanic or Latino (2135-2) | present/absent |  |
| Not Hispanic or Latino | Not Hispanic or Latino (2186-5) | present/absent |  |
| Unknown Ethnicity | UN | present/absent |  |
| Asian | Asian (2028-9) |  |  |
| Black or African American | Black or African American (2054-5) | present/absent |  |
| White | White (2106-3) | present/absent |  |
| Unknown Race | Unknown Race (2131-1) | present/absent |  |
| **Diagnosis** | **ICD-10 name (code)** |  |  |
| Heart diseases | Ischemic heart diseases (I20-I25) | present/absent | Anytime up to 1 day before Paxlovid treatment |
| Cancer | Neoplasms (C00-D49) | present/absent | Anytime up to 1 day before Paxlovid treatment |
| Hypertension | Essential (primary) hypertension (K10) | present/absent | Anytime up to 1 day before Paxlovid treatment |
| Cerebrovascular diseases | Cerebrovascular diseases (I60-I69) | present/absent | Anytime up to 1 day before Paxlovid treatment |
| Chronic lower respiratory diseases | Chronic lower respiratory diseases (J40-J47) | present/absent | Anytime up to 1 day before Paxlovid treatment |
| Chronic kidney diseases | Chronic kidney disease (CKD) (N18) | present/absent | Anytime up to 1 day before Paxlovid treatment |
| Liver diseases | Diseases of liver (K70-K77) | present/absent | Anytime up to 1 day before Paxlovid treatment |
| Overweight and obesity | Overweight and obesity (E66) | present/absent | Anytime up to 1 day before Paxlovid treatment |
| Type 2 diabetes | Type 2 diabetes mellitus (E11) | present/absent | Anytime up to 1 day before Paxlovid treatment |
| Disorders involving the immune mechanisms | Certain disorders involving the immune mechanism (D80-D89) | present/absent | Anytime up to 1 day before Paxlovid treatment |
| Congenital disorders | Congenital malformations, deformations and chromosomal abnormalities (Q00-Q99) | present/absent | Anytime up to 1 day before Paxlovid treatment |
| Mood disorders including depression | Mood [affective] disorders (F30-F39) | present/absent | Anytime up to 1 day before Paxlovid treatment |
| Psychotic disorders | Schizophrenia, schizotypal, delusional, and other non-mood psychotic disorders (F20-F29) | present/absent | Anytime up to 1 day before Paxlovid treatment |
| Behavioral disorders | Behavioral and emotional disorders with onset usually occurring in childhood and adolescence (F90-F98) | present/absent | Anytime up to 1 day before Paxlovid treatment |
| Substance use disorders | Mental and behavioral disorders due to psychoactive substance use (F10-F19) | present/absent | Anytime up to 1 day before Paxlovid treatment |
| Alzheimer’s disease | Alzheimer's disease (G20) | present/absent | Anytime up to 1 day before Paxlovid treatment |
| HIV | Human immunodeficiency virus [HIV] disease (B20) | present/absent | Anytime up to 1 day before Paxlovid treatment |
| Thalassemia | Thalassemia (D56) | present/absent | Anytime up to 1 day before Paxlovid treatment |
| Organ Transplant | Transplanted organ and tissue status (Z94) | present/absent | Anytime up to 1 day before Paxlovid treatment |
| Tobacco smoker | Nicotine dependence, unspecified, uncomplicated (F17.200) | present/absent | Anytime up to 1 day before Paxlovid treatment |
| **Medication and procedures** | RxNorm for medication coding  **Current Procedural Terminology (CPT)** for procedures |  | Anytime up to 1 day before Paxlovid treatment |
| Immunosuppressants | IMMUNOSUPPRESSANTS (L04) | present/absent | Anytime up to 1 day before Paxlovid treatment |
| COVID-19 Vaccination | - SARS-CoV-2 (COVID-19) Vaccine (213) - Severe acute respiratory syndrome coronavirus 2 (SARS-CoV-2) (Coronavirus disease [COVID-19]) vaccine, mRNA-LNP, spike protein, preservative free, 30 mcg/0.3mL dosage, diluent reconstituted, for intramuscular use (91300) - Severe acute respiratory syndrome coronavirus 2 (SARS-CoV-2) (Coronavirus disease [COVID-19]) vaccine, mRNA-LNP, spike protein, preservative free, 100 mcg/0.5mL dosage, for intramuscular use (91301) - Severe acute respiratory syndrome coronavirus 2 (SARS-CoV-2) (coronavirus disease [COVID-19]) vaccine, DNA, spike protein, chimpanzee adenovirus Oxford 1 (ChAdOx1) vector, preservative free, 5x10^10 viral particles/0.5mL dosage, for intramuscular use (91302) - Severe acute respiratory syndrome coronavirus 2 (SARS-CoV-2) (coronavirus disease [COVID-19]) vaccine, DNA, spike protein, adenovirus type 26 (Ad26) vector, preservative free, 5x10^10 viral particles/0.5mL dosage, for intramuscular use (91303) - Severe acute respiratory syndrome coronavirus 2 (SARS-CoV-2) (coronavirus disease [COVID-19]) vaccine, mRNA-LNP, spike protein, preservative free, 30 mcg/0.3 mL dosage, tris-sucrose formulation, for intramuscular use (91305) - Severe acute respiratory syndrome coronavirus 2 (SARS-CoV-2) (coronavirus disease [COVID-19]) vaccine, mRNA-LNP, spike protein, preservative free, 50 mcg/0.25 mL dosage, for intramuscular use (91306) - Severe acute respiratory syndrome coronavirus 2 (SARS-CoV-2) (coronavirus disease [COVID-19]) vaccine, mRNA-LNP, spike protein, preservative free, 10 mcg/0.2 mL dosage, diluent reconstituted, tris-sucrose formulation, for intramuscular use (91307) | present/absent | Anytime up to 1 day before Paxlovid treatment |

**eTable 2**: COVID-19 outcome measures, their standardized names, codes, data types and time window that are used in the TriNetX database.

| **Rebound outcomes** | **Name, code** | **Data type** | **Time Window** |
| --- | --- | --- | --- |
| COVID-19 infections | - COVID-19 (ICD-19 code U07.1) - SARS coronavirus 2 and related RNA [Presence] (labResult: Positive) (Laboratory code TNX:9088) | present/absent | 2-8 days after Paxlovid treatment |
| COVID-19 symptoms | - Disturbances of smell and taste (ICD-10 code:R43) - Fever of other and unknow origin (ICD-10 code: R50) - Headache (ICD-10 code: R51) - Pain (ICD-10 code: R52) - Malaise and fatique Fever of other and unknow origin (ICD-10 code: R53) - Cough (ICD-10 code: R05) - Shortness of breadth (ICD-10 code: R06.02) - Nausea and vomiting (ICD-10 code: R11) - Diarrhea, unspecified(ICD-10 code: R50) - Acuate pharyngitis (ICD-10 code: J02) - Nasal congestion (ICD-10 code: R09.81) - Abnormalities of breathing (ICD-10 code: R06) - Pain in the throat and chest (ICD-10 code: R07) - Disorientation, unspecified (ICD-10 code: R41.0) |  |  |

### Propensity Score Matching

Propensity score matching was performed on all listed characteristics in eTable 1. Characteristics of the cohorts before and after matching are summarized in the table below.

| **Cohort 1 and cohort 2 patient count before and after propensity score matching** | | | | | |
| --- | --- | --- | --- | --- | --- |
|  | | Cohort | Patient count before matching | | Patient count after matching |
|  | | 1 – BA.5 cohort | 5,161 | | 5,142 |
|  | | 2 - BA.2.12.1 cohort | 10,752 | | 5,142 |
| **Propensity score density function - Before and after matching (cohort 1 - purple, cohort 2 - green)** | | | | | |
|  |  | 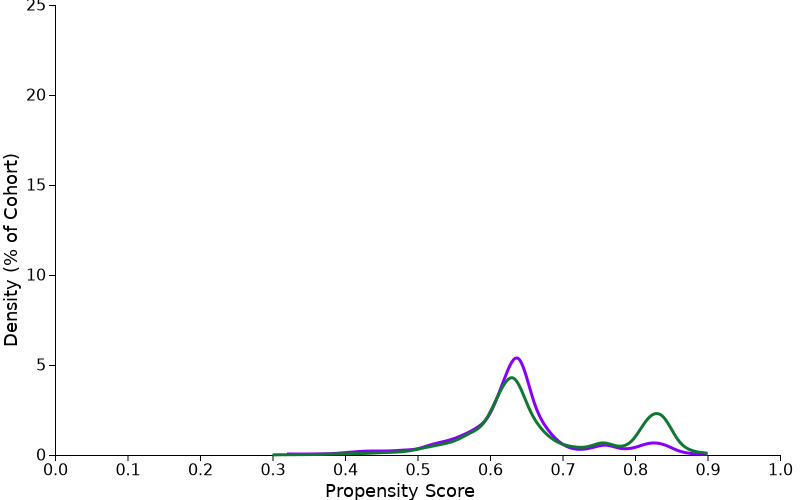 | | 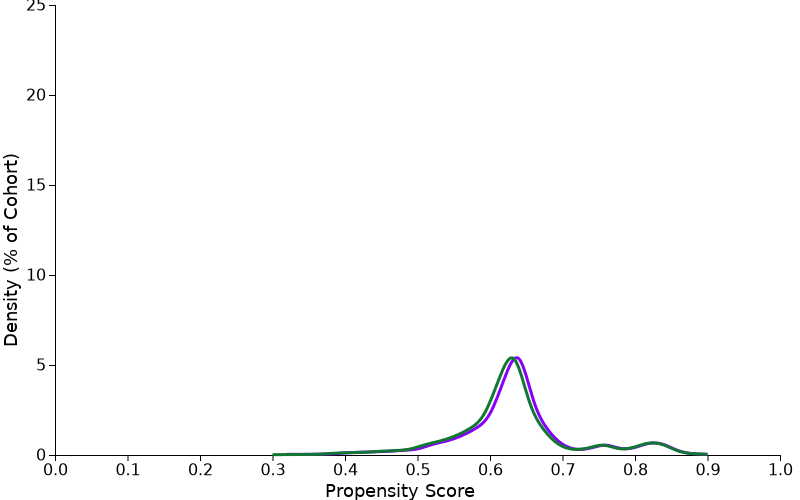 | |

**Statistical methods:**

The Kaplan-Meier Analysis estimates probability of the outcome at a respective time interval (daily time interval is used in this analysis). In order to account for the patients who exited the cohort during the analysis period, and therefore should not be included in the analysis, censoring is applied. In this analysis, patients are removed from the analysis (censored) after the last fact in their record.

The proportional hazard assumption was tested using the generalized Schoenfeld approach. The TriNetX Platform calculates the hazard ratios and associated confidence intervals using R's Survival package v3.2-3. For generating hazard ratios, TriNetX sets robust=FALSE using the R survival package, which is a limitation of the TriNetX platform since it does not consider potential clustering of COVID-19 cases within the healthcare organizations or specific geolocations. All Statistical tests were conducted on 8/4/2022 within the TriNetX Analytics Platform with significance set at p-value < 0.05 (two-sided).
